# Operation Theatre cancellations highly associated with poor OT time management: 8 month cross-sectional study at a tertiary center in Ethiopia

**DOI:** 10.1101/2025.08.01.25332783

**Authors:** Gersam A Mulugeta, Kumlae T Daba, Lidya G Mude, Birhanu A Tesso

**Affiliations:** Department of Surgery, Jimma University, Jimma, Ethiopia; Department of Ophthalmology, Jimma University, Jimma, Ethiopia

**Keywords:** Cancellation, First case incision time, OR efficiency, OR utilization, Reasons

## Abstract

**Background:** Improving Operating Theater (OT) efficiency enhances productivity, reduces patient waiting time, and boosts satisfaction. First case start time determines operating table utilization, making the first patient the “golden patient.” Delays in first case incision time can lead to case cancellations. This study aimed to identify factors affecting first elective case incision time (FCIT) and operating room (OR) cancellations at Jimma University Medical Center (JUMC).

**Objective:** To assess the prevalence and causes of OR cancellation and delayed first case start in elective major surgical procedures at JUMC between September 2022 and April 2023.

**Methods:** A prospective cross-sectional study was conducted on elective surgeries in four OTs at JUMC. Data on patient entry (wheels-in), anesthesia initiation, incision time, anesthesia completion, and patient exit (wheels-out) were recorded. Factors like hospital stay, consent status, and pre-anesthetic evaluations were also documented. Data were collected by trained nurses and analyzed using SPSS version 23 and Excel.

**Results & Discussion:** Of 1,292 scheduled elective surgeries (M:F = 1:1.2, mean age 29 ± 22 years), 519 (40.2%) were planned first cases, out of which 479 (91.6%) were delayed. Key delay reasons were OR material preparation (53%), staff issues (15.3%), and patient condition (5.6%). The surgical cancellation rate was 20%, with time insufficiency (41%) and patient-related conditions (28%) as main causes. Consent was unsigned in 12% of cases. The average wheels-in time was 8:18 AM, with anesthesia induction at 8:38 AM. OR utilization was 1.8 cases/day, below the recommended 3 cases/day.

**Conclusion:** Most delays and cancellations were modifiable. A quality improvement (QI) project focused on timeliness with planned sustainability could enhance OR efficiency.

## Background

The World Health Organization (WHO), in collaboration with the Organisation for Economic Co-operation and Development (OECD) and The World Bank, published the report *Delivering Quality Health Services: A Global Imperative for Universal Health Coverage* in 2018. This report underscores the crucial importance of delivering quality health services as a core pillar of achieving Universal Health Coverage (UHC) (1). Universal Health Coverage aims to ensure that all individuals, regardless of their socioeconomic status, geographic location, or other factors, have access to the necessary healthcare services without financial hardship. The WHO, OECD, and World Bank emphasize that UHC is not merely about the availability of healthcare services but also about ensuring that these services are effective, equitable, and of high quality (1).

Due to the pressure in health from requirements of aging population, introduction of expensive medical technologies and greater community expectations for access to health services, public hospitals are required to provide better care by being more efficient and reducing wasteful spending. Efficiency improvement in the OT can improve OT productivity, decrease patient waiting time to surgery and increase patient satisfaction. Numerous factors in the perioperative setting influence OT productivity, such as surgical scheduling accuracy, starting on time, minimising procedure time variation (but taking in procedure complexity), turnover time, inter-operative delays and bed management. In addition, the training of junior surgical and anesthetic staff may affect OT productivity (2,3).

First case starting time determines the utilization of the operating table and thus the first patient is termed the golden patient(4). Multiple factors related to HR, equipment, consumables and others can delay the first case incision time. This intern may lead to cancellation of the last cases. Cancellation may also arise from poor pre-operative preparation, unforeseen infrastructural challenges, or over planning. As we are living in a resource limited country, efficient utilization of the operating tables are paramount. Range of reasons for Cancellation of elective scheduled operations include inadequate pre-op assessment and preparation, patient-related factors, lack of operating room time, Improper work-up, Facility related, and surgeon-related issues (3,5).

Improving the efficiency of surgery was the first objective of the 6th strategic pillar of The Safe Surgery 2020 (SS2020) project which ended in 2020. OT efficiency indicators include Rate of First Elective Case On-Time Theater Performance and Rate of Cancellation of Elective Surgery. Majority of the high income countries have OR cancellation rates ranging between 0.88% and 10% while low income countries have as high as 30%. This study aimed to identify factors affecting the first elective case On-time (First case incision time – FCIT) and OR cancellation at Jimma University Medical Center, a tertiary teaching hospital in LMIC. A consequent quality improvement implementation study will be conducted to address the most prevalent factors. (6)

### Objectives

The aim of this study was to assess the prevalence and causes of operating room (OR) cancellation rates and delayed first surgery start times in elective surgical patients scheduled for major surgical procedures at Jimma University Medical Center (JUMC) between September 2022 and April 2023. By identifying the factors contributing to cancellations and delays, the study sought to provide insights into the operational challenges and areas for improvement in the surgical scheduling process at JUMC.

## Methods

### Study Site

The study was conducted at Jimma University Medical Center (JUMC), located 360 km southwest of Addis Ababa. JUMC is a tertiary referral hospital serving a catchment population of approximately 15 million people. It has an 800-bed capacity and operates 8 surgical theaters.

### Study Period

The study was carried out from September 2022 to April 2023.

### Study Design

A cross-sectional prospective study design was employed.

### Study Subjects

The study included all patients scheduled for elective day-case surgeries in operating rooms 2, 3, 4, 6, and 8 during the study period.

### Variables

The study variables included

- **Primary outcome variables**:
  - Operating room (OR) cancellation rate and reasons for cancellation.
  - First-case surgery start time and reasons for delay beyond the hospital-agreed time of 8:30 AM.
- **Independent variables**:
  - OR times: Wheels-in time, anesthesia start time, surgery incision time, anesthesia end time, and wheels-out time.
  - Preoperative factors:
    - Duration of hospital stay before surgery.
    - Consent status at the OR door.
    - Attachment of the pre-anesthetic evaluation sheet at the OR door.

### Data Collection Process

Data were collected prospectively by trained personnel using a structured data collection sheet. OR staff and anesthetists recorded the exact timing of key surgical events, including patient entry into the OR (wheels-in), anesthesia initiation, surgical incision time, completion of anesthesia, and patient exit from the OR (wheels-out). Additionally, information regarding cancellations and the reasons for delays in first-case surgeries was documented. Preoperative factors, such as the patient’s hospital stay before surgery, consent status, and availability of pre-anesthetic evaluation sheets, were also recorded at the time of surgery.

### Data Quality control

The data quality control was ensured with the following measures: The data collectors underwent a one-day training session. A pilot study was conducted over one week, after which the questionnaire was evaluated and refined accordingly. The data collection process was monitored on a weekly basis. Before entry into the Kobo Toolbox for analysis, the collected data were reviewed for completeness.

### Data Analysis

The collected data were entered into Kobo Toolbox and exported to SPSS version 27 and Microsoft Excel sheet version 2013 for analysis. Descriptive statistics, including frequencies and percentages, were used to summarize categorical variables, while means and standard deviations were calculated for continuous variables. The prevalence of OR cancellations and delayed first-case start times was determined, and factors contributing to these issues were analyzed.

### Ethical Considerations

Ethical clearance was obtained from the Institutional Review Board (IRB) of Jimma University (IRB approval number – IHRPGI441/22). Permission to conduct the study was granted by the hospital administration. The study was conducted in accordance with the ethical principles outlined in the Declaration of Helsinki. Patient confidentiality was strictly maintained, and no personally identifiable information was collected.

## Results

A total of 1,292 surgeries were scheduled for elective procedures, with a male-to-female ratio of 1:1.2. The patients’ ages ranged from 1 month to 85 years, with a mean age of 29 ± 22 years. The average duration of hospital stay before surgery was 7.1 days (median = 5 days), with a range of 1 to 90 days. The majority of patients (511, 39.6%) were admitted to the general surgical ward, followed by the pediatric surgical ward (386, 29.9%) and the gynecology ward (204, 15.8%) (See Table 1).

**Table 1:** Admission ward and operation room of scheduled patients for elective surgery at Jimma University Medical center, Sept-Feb 2022.

| Case operated | Operation room | Number | Percent (%) |
| --- | --- | --- | --- |
| **General Surgery** | Room 3, 4 | 511 | 39.6 |
| **GI/ Hepatobiliary** | Room 8 | 158 | 12.2 |
| **Gynecology** | Room 3, 4 | 204 | 15.8 |
| **Neuro surgery** | Room 2, 6 | 7 | 0.5 |
| **Ophthalmology** | Room 2 | 12 | 0.9 |
| **Pediatrics Surgery** | Room 2 | 386 | 29.9 |
| **Urology** | Room 6 | 14 | 1.1 |
| **Total** |  | 1292 | 100 |

### Surgical Case Distribution and First-Case Incision Time

Among the scheduled cases, 519 (40.2%) were planned as first cases (See figure 1). The total number of first cases that experienced delays were 479 (91.6%). The median FCIT for the 6 month period ranged from 8:30AM – 9:20AM (i.e. 50 minutes late from the agreed start time). The run chart below (see figure 2) shows improvement in the middle of the study period between October and November while there was another project running in the hospital which involved intense administrative follow up of the operation theatre time. The follow up did not continue beyond November after which the data shows that the incision time gradually worsened back. The main reason for delay to start surgery was OR material preparation (142 (53%)) followed by staff issues (41(15.3%)) and patient condition (15(5.6%)).

**Figure 1:**
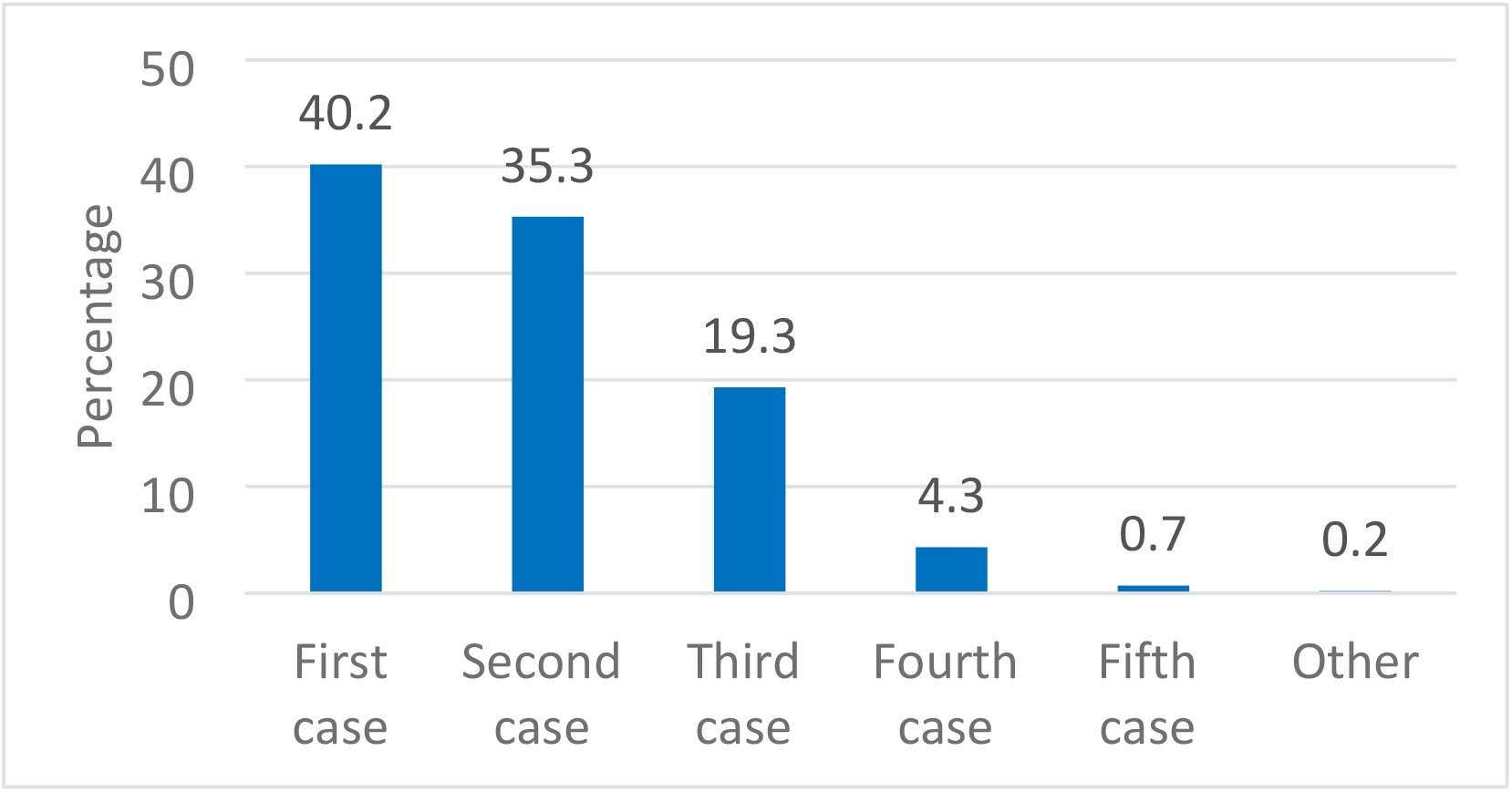
Surgical case distribution by the order of operation at Jimma University Medical center, Sept-Feb 2022

**Figure 2:**
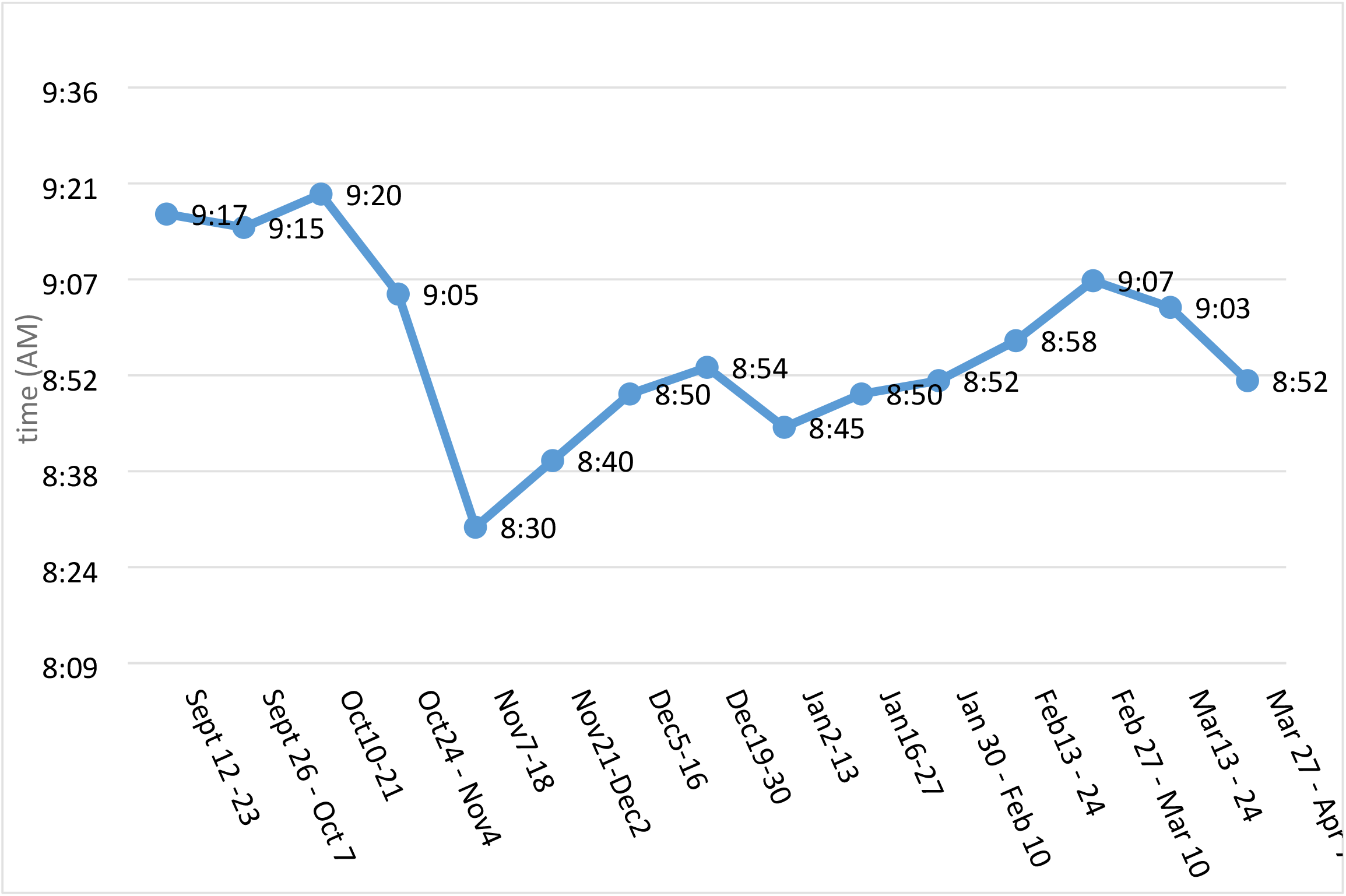
Median First Case Incision Time at Jimma University Medical center, Sept-Feb 2022

### Surgical Cancellation Rate and Contributing Factors

The average surgical cancellation rate during the study was 20%. Initially, the cancellation rate was as high as 32% but was significantly reduced to 20% after six months of project implementation. During the period from January 2 to 13, the cancellation rate was 9%, largely due to the fact that fewer surgeries were performed as a result of anesthesia drug shortages. Those that were scheduled were few and they did not get canceled. (See figure 3).

**Figure 3:**
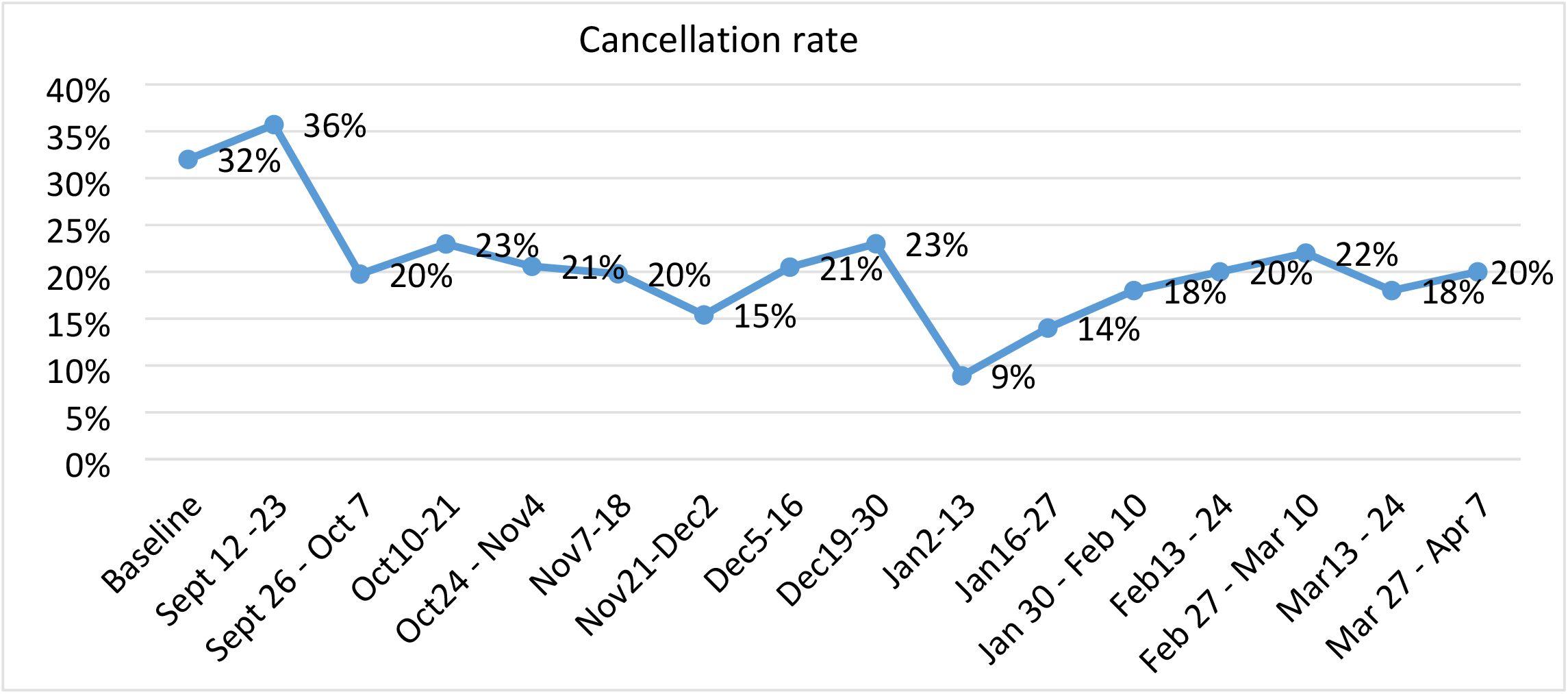
Surgical cancellation rate at Jimma University Medical center, Sept-Feb 2022

The main reasons for surgery cancellations included (See figure 4):

**Figure 4:**
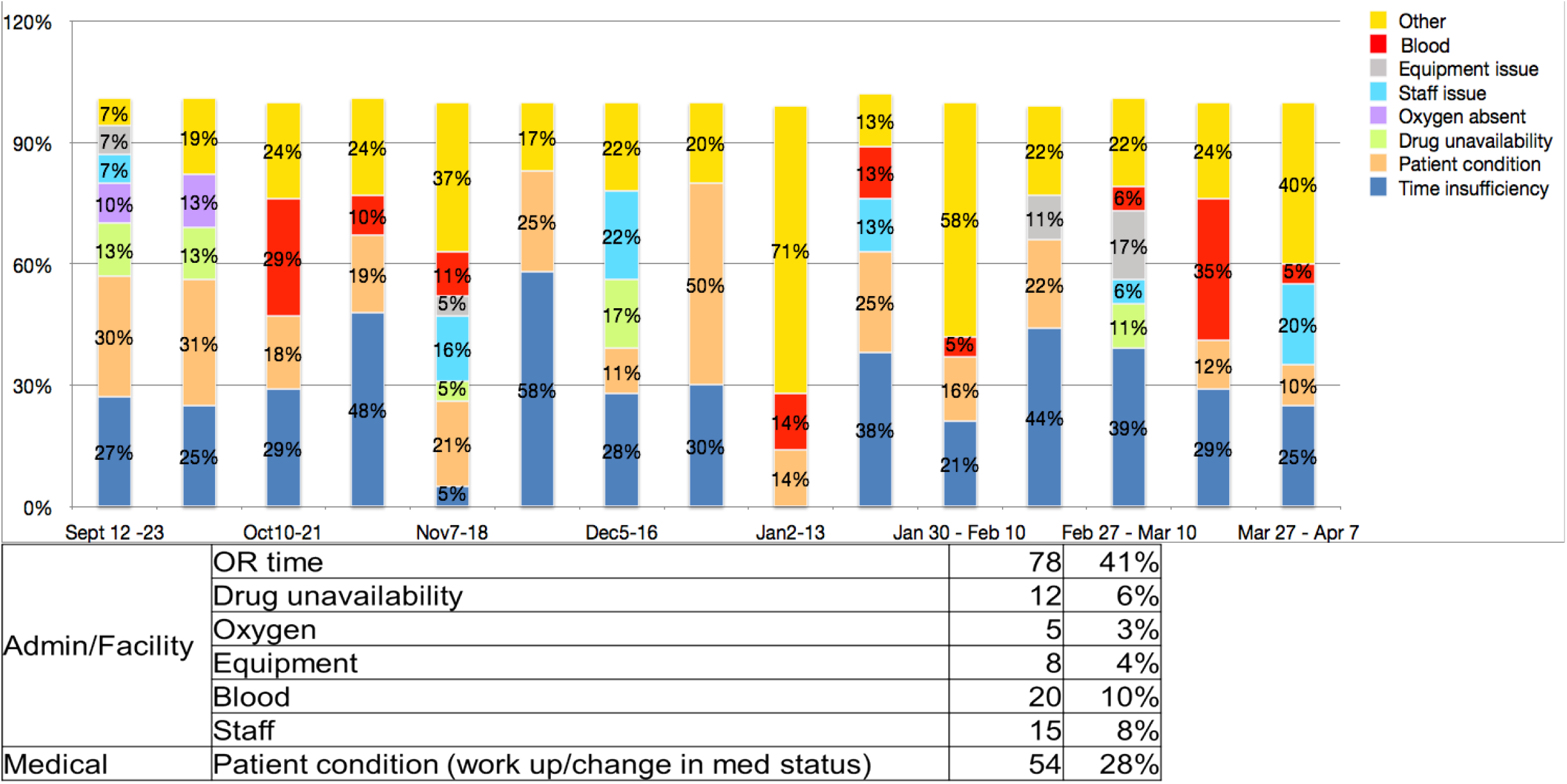
Reasons for surgical cancellation rate at Jimma University Medical center, Sept-April 2022

- **Time insufficiency** (78 cases, 41%)
- **Patient-related conditions** (54 cases, 28%)
- **Other factors**, including a shortage of blood, staff issues, drug unavailability, equipment malfunction, and oxygen shortages

### Operating Room Efficiency Metrics

- **Wheels-in time**: The average time patients entered the OR was 8:18 AM.
- **Anesthesia induction time**: The average anesthesia induction time was 8:39 AM, improving from 8:55 AM at the start of the project.
- **Surgery start time after anesthesia induction**: The average interval between anesthesia induction and surgery start was 21 minutes, improving from 25 minutes to 18 minutes by the end of the study.
- **Surgery duration**: The average surgical duration was 1 hour and 50 minutes.
- **Time from surgery end to wheels-out**: The average time patients remained in the OR after surgery completion was 20 minutes, improving from 24 minutes at the start of the project to 17 minutes by the end.
- **Overall operating theater utilization**: The average OR utilization rate was 1.8 surgeries per day per room.

The study found that consent was not signed at the time the patient arrived at the operating room (OR) door in 12% of cases, while the pre-anesthetic evaluation sheet was attached in 59% of cases. The average wheels-in time was recorded at 8:18 AM, with anesthesia induction occurring at an average of 8:38 AM. The average duration of surgery was 1 hour and 50 minutes, while overall operating theater utilization stood at 1.8. Additionally, patients had an average hospital stay of seven days before undergoing elective surgery, indicating potential areas for improving preoperative efficiency and patient flow.

## Discussion

The patients’ ages ranged from 1 month to 85 years, with a mean age of 29 ± 22 years. This broad age distribution highlights the diverse patient population undergoing elective surgeries.

Out of 1,292 planned surgical cases, 1,036 patients underwent surgery, resulting in a cancellation rate of 19.8%. This rate is comparable to many centers in low- and middle-income countries (LMICs) but remains higher than those reported in high-income countries (HICs). In LMIC settings, reported cancellation rates include 24.5% at Tikur Anbessa Specialized Hospital (TASH) in Ethiopia, 22.4% in Wolayta, 20% in Sudan, and 28.8% in Uganda (7–10). In contrast, HIC institutions report significantly lower rates, such as 6.6% at the University of Pennsylvania (USA), 7.6% in Hong Kong, 6.79% in Brazil, and just 0.88% in India (11–14). Notably, some LMIC centers, such as St. Paul’s Hospital in Ethiopia (8.9%) and institutions in Nigeria (9.1%), report lower cancellation rates, highlighting the potential for improvement in surgical efficiency within similar resource-limited settings (15,16).

Among the 519 cases scheduled as first-case surgeries, 467 were successfully performed, yielding an 11% cancellation rate. However, only 40 cases (8.6%) commenced on time at 8:30 AM. When considering a 10-minute grace period (allowing surgeries to begin by 8:40 AM), the number of on-time cases increased to 104 (22.3%). These findings highlight persistent delays in first-case start times, emphasizing the need for targeted interventions to improve OR efficiency and scheduling adherence.

The primary causes of surgical delays were related to operating room (OR) preparation (53%), followed by staff-related issues (15.3%) and patient conditions (5.6%). Equipment malfunctions accounted for 4.1% of the delays, while anesthesia drug shortages contributed to 1.9%. Addressing these factors through strategic interventions could enhance surgical start times and overall OR efficiency.

These findings align with other studies. In the United States, delays were primarily attributed to surgeons (56.5%), preoperative processes (18.3%), and room-related factors (13%) (17). A multicenter study in Ethiopia identified key contributing factors such as patient late arrival to the OR (2.5 times increased likelihood), abnormal laboratory results (2.4 times), and surgeon late arrival (10 times) (18). These comparisons underscore the widespread nature of surgical delays and the influence of multiple modifiable factors.

A separate local quality improvement project was implemented between October and November, involving strict follow-up by high-level officials on OR start times. During this period, the average first-case incision time (FCIT) improved significantly to 8:30AM but the delay worsened back in the subsequent months making the average FCIT 9:01AM, mean delay of 29 minutes. This reinforces the need for sustained interventions to improve OR efficiency and adherence to scheduled start times.

## Conclusion and recommendation

The study found a high surgical cancellation rate of 19.8%, with only 8.6% of first-case surgeries starting on time. Operating room (OR) time inadequacy was the leading cause of cancellations, accounting for 57%. Importantly, all identified reasons for OR cancellations and first-case surgery delays are modifiable. Implementing a quality improvement (QI) project with a focus on sustainability could lead to significant improvements in OR efficiency and timely surgical procedures.

## Data Availability

Data is available with the corresponding author and can be made available upon request.

